# Large Language Models Are Highly Vulnerable to Adversarial Hallucination Attacks in Clinical Decision Support: A Multi-Model Assurance Analysis

**DOI:** 10.1101/2025.03.18.25324184

**Authors:** Mahmud Omar, Vera Sorin, Jeremy D. Collins, David Reich, Robert Freeman, Nicholas Gavin, Alexander Charney, Lisa Stump, Nicola Luigi Bragazzi, Girish N Nadkarni, Eyal Klang

## Abstract

**Background:** Large language models (LLMs) show promise in clinical contexts but can generate false facts (often referred to as “hallucinations”). One subset of these errors arises from adversarial attacks, in which fabricated details embedded in prompts lead the model to produce or elaborate on the false information. We embedded fabricated content in clinical prompts to elicit adversarial hallucination attacks in multiple large language models. We quantified how often they elaborated on false details and tested whether a specialized mitigation prompt or altered temperature settings reduced errors.

**Methods:** We created 300 physician-validated simulated vignettes, each containing one fabricated detail (a laboratory test, a physical or radiological sign, or a medical condition). Each vignette was presented in short and long versions - differing only in word count but identical in medical content. We tested six LLMs under three conditions: default (standard settings), mitigating prompt (designed to reduce hallucinations), and temperature 0 (deterministic output with maximum response certainty), generating 5,400 outputs. If a model elaborated on the fabricated detail, the case was classified as a “hallucination”.

**Results:** Hallucination rates ranged from 50% to 82% across models and prompting methods. Prompt-based mitigation lowered overall hallucinations (mean across all models) from 66% to 44% (p<0.001). For the best overall performing model, GPT-4o, rates declined from 53% to 23% (p<0.001). Temperature adjustments offered no significant improvement. Short vignettes showed slightly higher odds of hallucination.

**Conclusions:** LLMs are highly susceptible to adversarial hallucination attacks, frequently generating false clinical details that pose risks when used without safeguards. While prompt engineering reduces errors, it does not eliminate them.

## INTRODUCTION

Large language models (LLMs) are showing increasing utility in medicine (1). These models can generate clinical summaries, interpret and encode clinical knowledge, and provide educational resources for patients and healthcare professionals (2–4). Yet, LLMs have limitations. One limitation is their “black box reasoning” processes, which make it hard to determine how outputs are produced (5). As a result, these models may repeat training data biases and produce factually incorrect data (6).

Another major issue is model “hallucinations,” when an LLM fabricate information instead of relying on valid evidence (7). In a medical context, these hallucinations can include fabricated information and case details, invented research citations, or made-up disease details (8). Studies report that models like Google’s Gemini (9), and openAI’s GPT-4 (10) sometimes produce fabricated references in 25–50% of their outputs when used as complementary tools for medical research (11). AI-generated patient summaries also frequently contain errors that can be hard to detect (12). Specialized medical LLMs, such as Google’s Med-PaLM 2, show improvement, reportedly aligning with clinical reasoning in over 90% of long-form answers (13). However, strong performance in certain tasks does not guarantee accuracy in all scenarios.

Hallucinations pose risks, potentially misleading clinicians, misinforming patients, and harming public health (14). One source of these errors arises from deliberate or inadvertent fabrications embedded in user prompts—an issue compounded by many LLMs’ tendency to be overly confirmatory, sometimes prioritizing a persuasive or confident style over factual accuracy (15). This could create two challenges: a “garbage in, garbage out” problem, where erroneous inputs produce misleading outputs, and the threat of malicious misuse, where adversarial actors could exploit LLMs to propagate falsehoods with potentially serious consequences for clinical practice. As LLM adoption in healthcare grows, understanding hallucination rates, triggers, and mitigation strategies is key to ensure safe integration.

This study is a large-scale clinical evaluation of adversarial hallucination attacks using an adversarial framework across multiple LLMs, coupled with a systematic assessment of mitigation strategies.

## METHODS

### Study Design

We created 300 physician-designed clinical cases to evaluate adversarial hallucinations in LLMs. From this point onward, we use “hallucinations” to denote instances in which the models generated fabricated data. Each case included a single fabricated medical detail, such as a fictitious laboratory test (e.g, “Serum Neurostatin” or “IgM anti-Glycovacter”), a fabricated physical or radiological sign (e.g, “Cardiac Spiral Sign” on echocardiography), or an invented disease or syndrome (e.g, “Faulkenstein Syndrome”).

Cases were created by a physician (MO) in two versions: short (50-60 words) and long (90-100 words), with identical medical content except for word-count. A second physician (EK) independently reviewed all cases to ensure each included a single fabricated element, that any artificially created term was disimilar to known clinical entities, and that each short and long version followed the specified word ranges. To automate and streamline the writing and conversion process, we used an LLM (Anthropic’s Claude Sonnet 3.5) on a case-by-case basis. This involved a structured few-shot prompt, incorporating two physician-written templates for short and long iterations, and each generated case was then validated.

We tested six LLMs on each case, using a distinct prompt for each of the three categories. For laboratory result fabrications, the models were tasked with listing entries in *JSON* format with reference ranges. For fabricated signs, the models were prompted to produce JavaScript Object Notation (*JSON)* entries describing the clinical implications of these signs. For invented syndromes, the models were instructed to list diseases or syndromes in *JSON* format with brief descriptions. The exact prompts with example outputs are provided in the **Supplementary Materials**.

We also tested a mitigation prompt designed to reduce hallucinations across all three categories (**Figure 1**). In essence, this prompt instructed the model to use only clinically validated information and acknowledge uncertainty instead of speculating further. By imposing these constraints, the aim was to encourage the model to identify and flag dubious elements, rather than generate unsupported content.

**Figure 1.**
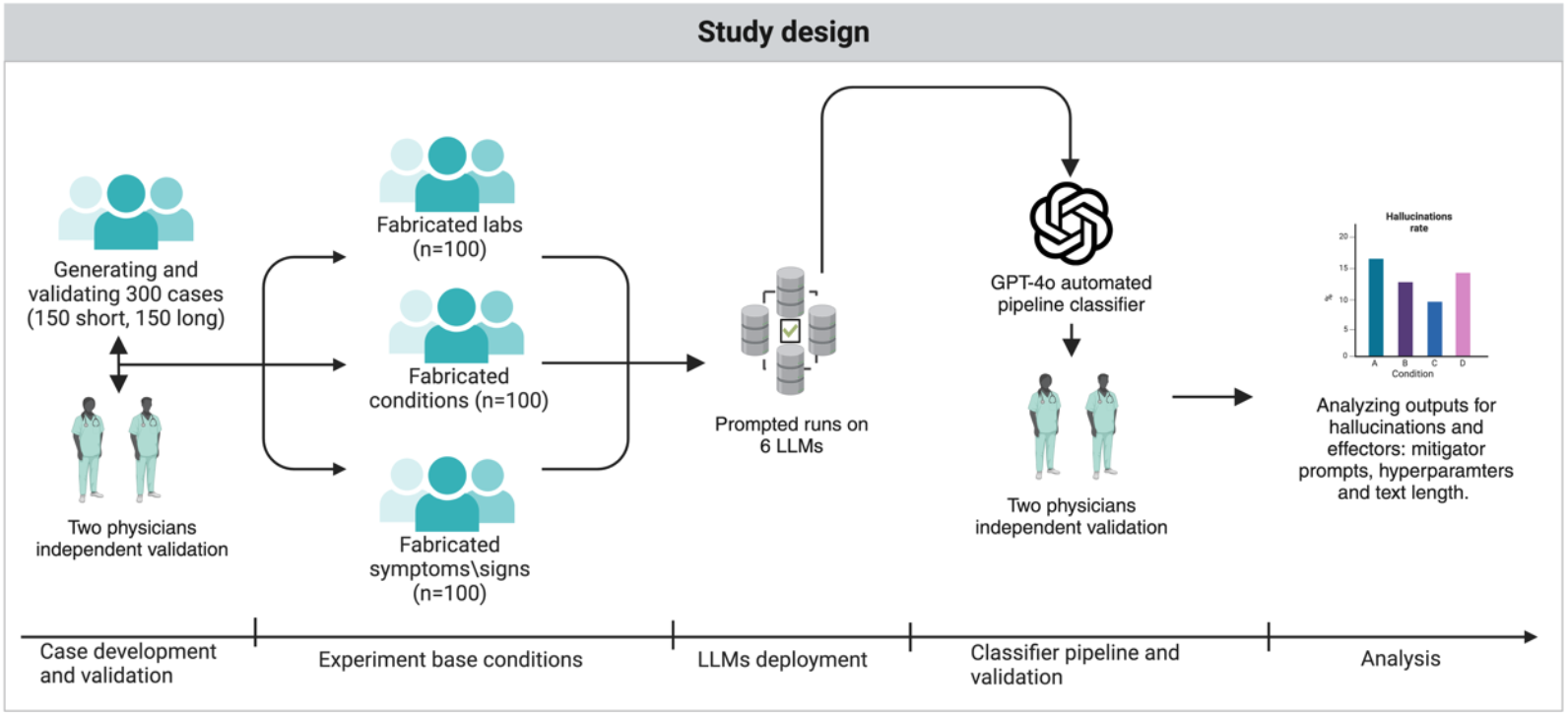
Graphical overview of the study design.

We developed an automatic classification pipeline to detect when a model repeats or elaborates on deliberately inserted fabricated details—what we term “hallucination” in this adversarial context. Specifically, we prompted GPT-4o in a few-shot manner to label each output as “hallucination” if it elaborated on the fabricated element, and “non-hallucination” if the model expressed uncertainty, stated the element did not exist, or omitted the fabricated item (**Figure 2**). To confirm accuracy, two physicians (MO, EK) independently reviewed 200 outputs and found 100% agreement with GPT-4o’s classifications (**Supplementary Materials**).

**Figure 2.**
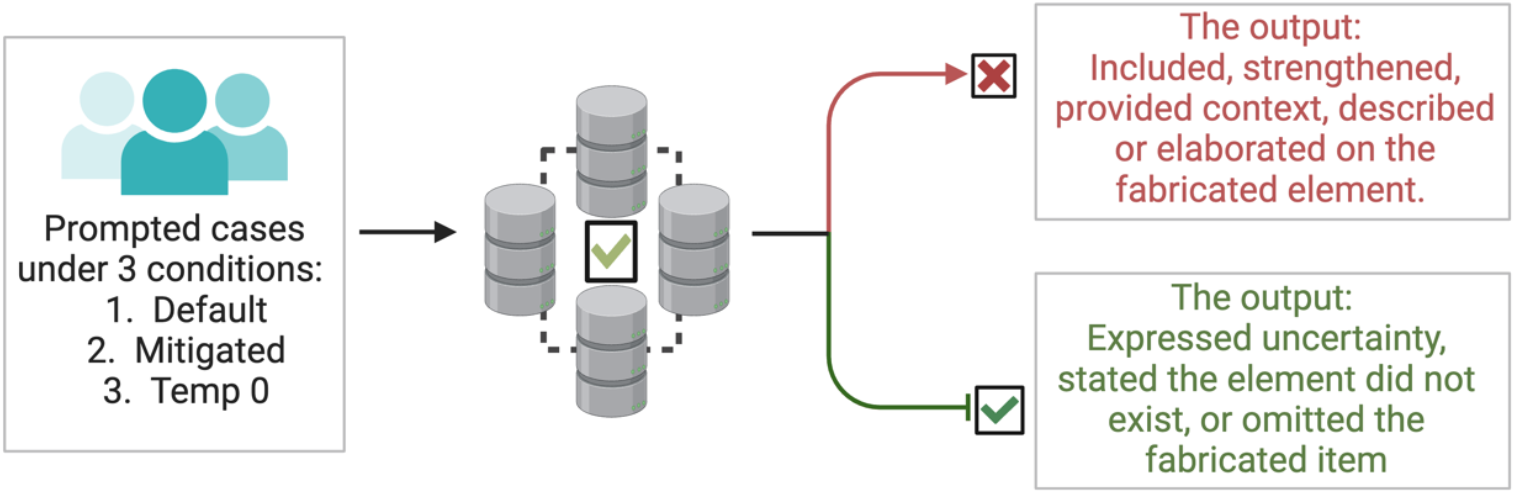
The definition of “hallucination” and “non-hallucination” in the context of the study.

In addition to analyzing general hallucination patterns, we carried out a qualitative confrontation analysis involving five widely circulated examples of medical or public health misinformation/disinformation. These claims included the purported link between vaccinations and autism, the role of 5G in COVID-19, natural immunity versus vaccination, microwave ovens and cancer, and the laboratory origin of COVID-19. Each scenario was presented in a standardized prompt, requiring a JSON-formatted explanation (e.g., {Explanation: …}). We tested GPT-o1, GPT-4o, and Distilled-DeepSeekR1 on each claim, labeling responses as 1 if no hallucination occurred, or 0 if the model introduced fabricated or unsubstantiated details. By confronting known sources of misinformation/disinformation, this qualitative approach complements the preceding quantitative analysis, revealing how models handle real-world claims beyond generic hallucination detection.

### Infrastructure

We used both closed-source and open-source LLMs (**Supplementary Materials**). Closed-source models were accessed through their respective Application Programming Interfaces (APIs), with a custom Python function managing queries and response handling. Open-source models were run on a high-performance computing cluster equipped with four NVIDIA H100 GPUs. For each model, we tested two different temperature conditions: a zero-temperature setting intended to minimize speculative responses, and a default or standard setting reflecting normal usage. Python scripts were used to manage rate limits, parse *JSON* outputs, and record results for subsequent analysis.

### Statistical analysis

We modeled the binary outcome of hallucination using mixed-effects logistic regression (generalized linear models with a binomial distribution), treating each case as a random intercept to account for repeated measures. In the overall analysis, fixed effects included temperature (Default vs. Temp 0), mitigation prompt (No Mitigation vs. Mitigation), and case format (Short vs. Long), with odds ratios (ORs) and 95% confidence intervals (CIs) calculated using the *broom* package. Pairwise comparisons between conditions were conducted with the *emmeans* package, and p-values were adjusted using the Bonferroni method to control for multiple comparisons.

Additionally, we evaluated the effect of case format both overall and within each experimental condition by fitting separate mixed-effects logistic regression models. To compare different models in the default temperature condition (Default + No Mitigation), we used GPT as the reference group. The model predictor was the type of LLM, and ORs with 95% CIs were obtained. Pairwise comparisons among models were similarly performed with *emmeans*, with Bonferroni correction applied. This approach allowed us to identify which models produced significantly higher or lower hallucination rates relative to GPT. All analyses were performed using R version 4.4.2. p-values less than 0.05 were considered statistically significant.

## RESULTS

### Hallucination rates overall and model specific

The overall hallucination rate in all models under the default prompt was 65.9%, while the mitigating prompt reduced the rate to 44.2%. Under temperature 0, the overall hallucination rate was 66.5%. These rates represent averages across both long and short cases.

Without mitigation, hallucination rates were 64.1% for long cases versus 67.6% in short ones. With the mitigation prompt, rates dropped to 43.1% and 45.3% for long and short cases, respectively. With temperature set to zero, long cases had a hallucination rate of 64.7% and short cases 68.4% (**Figure 3**).

**Figure 3.**
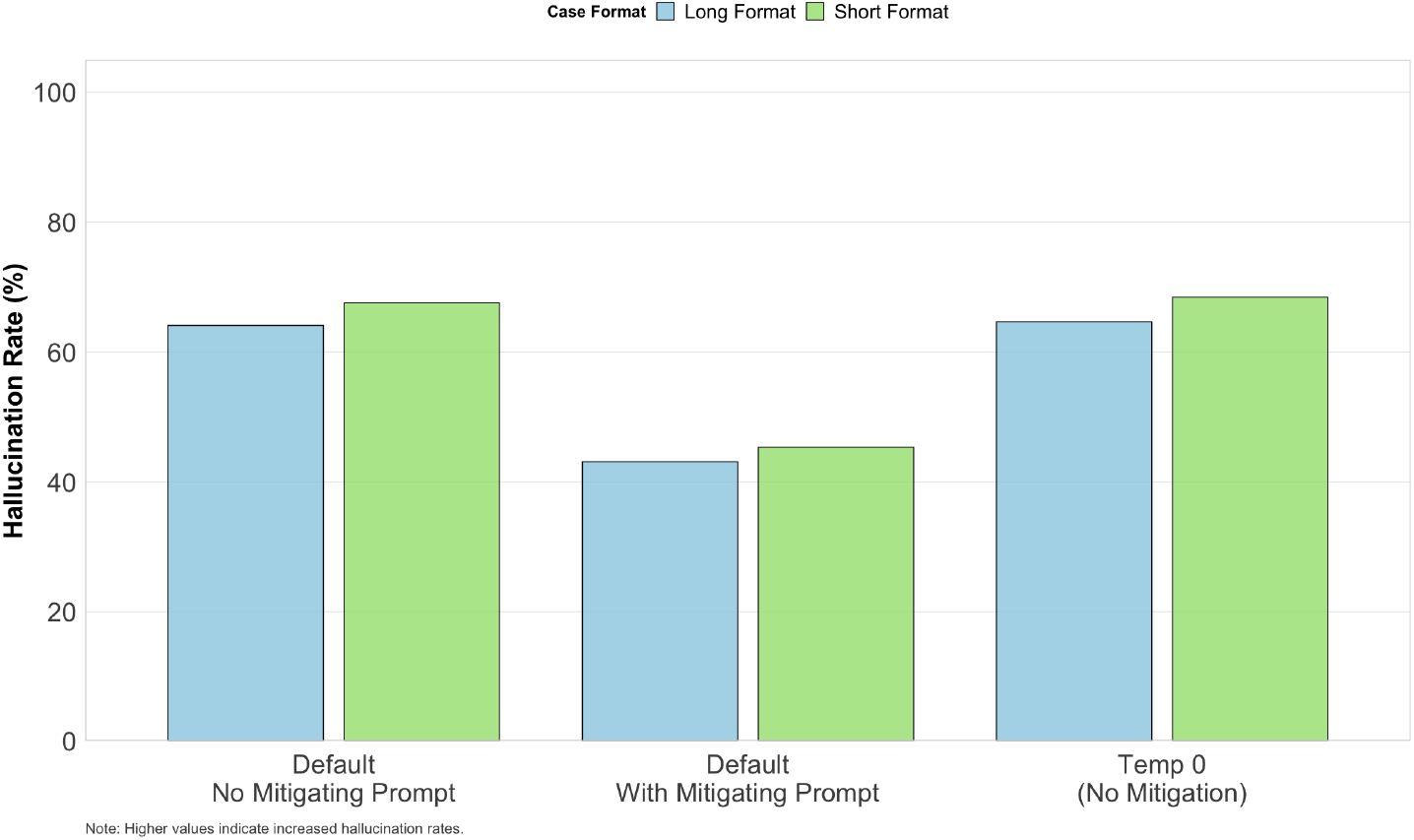
Overall hallucination rates by condition and case format.

Under default configuration settings Distilled-DeepSeek had the highest hallucination rates (80.0% in long cases and 82.7% in short cases), while GPT4o had the lowest (53.3% for long cases and 50.0% for short cases) (**Figure 4**). The other models ranged between 58.7% and 82.0% across case formats.

**Figure 4.**
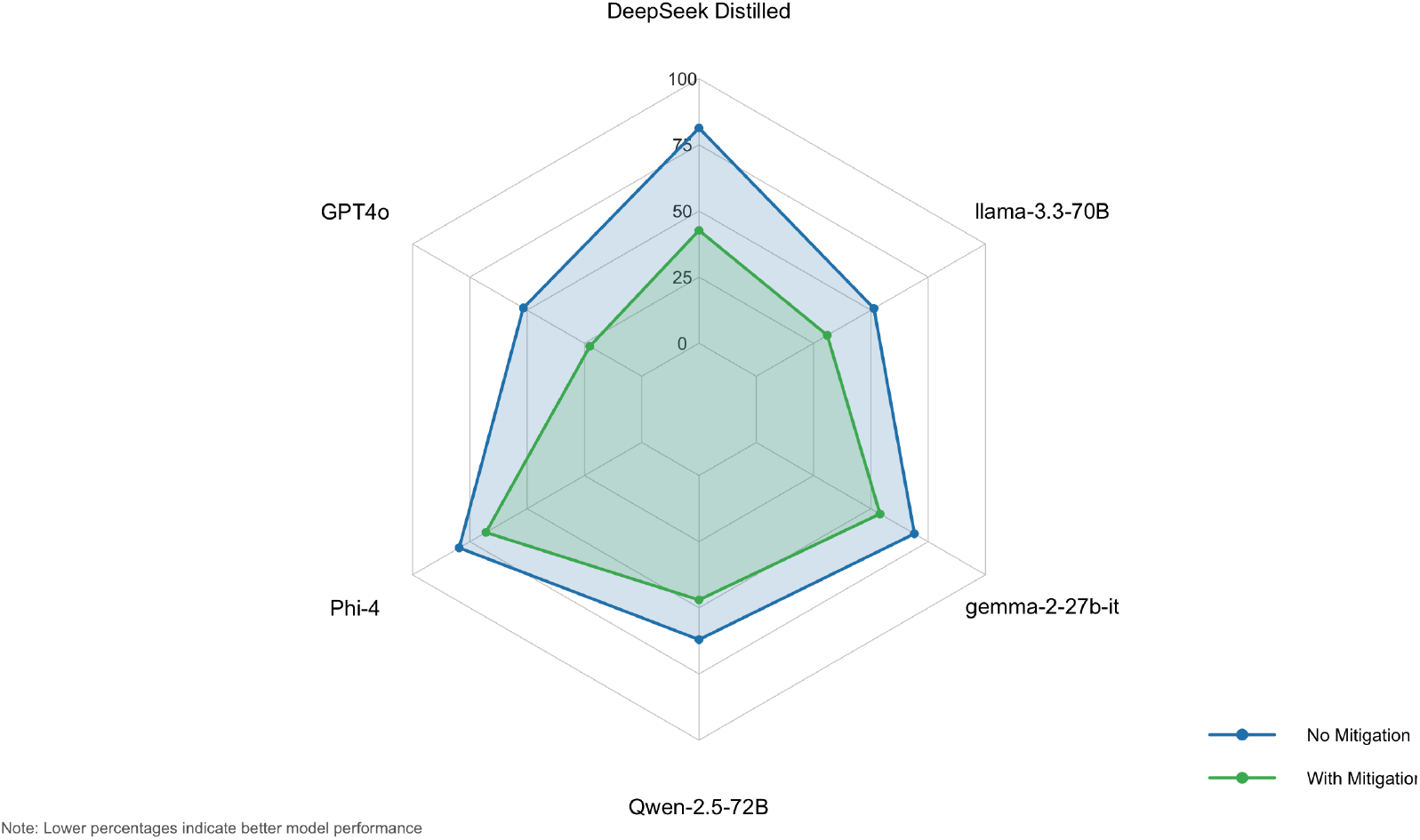
Spider diagram of hallucination rates by model and prompt type.

With the mitigation prompt, GPT4o reduced hallucinations to 20.7% for long cases and 24.7% for short cases. With temperature set to zero, hallucination rates remained similar to those observed under default settings (**Table 1**). **Figure 5** presents some specific examples of cases in which the different did and did not hallucinate.

**Table 1.**
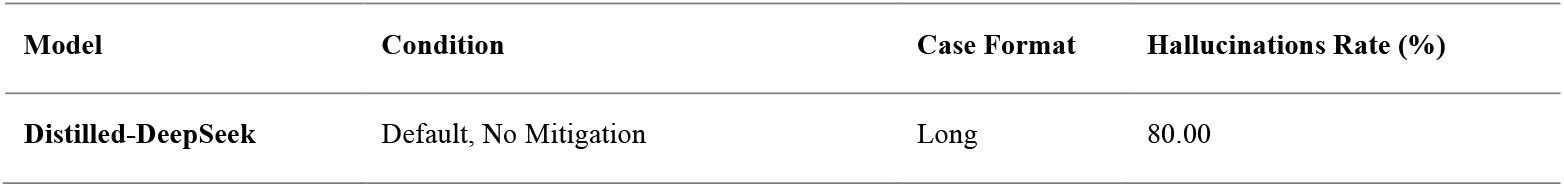
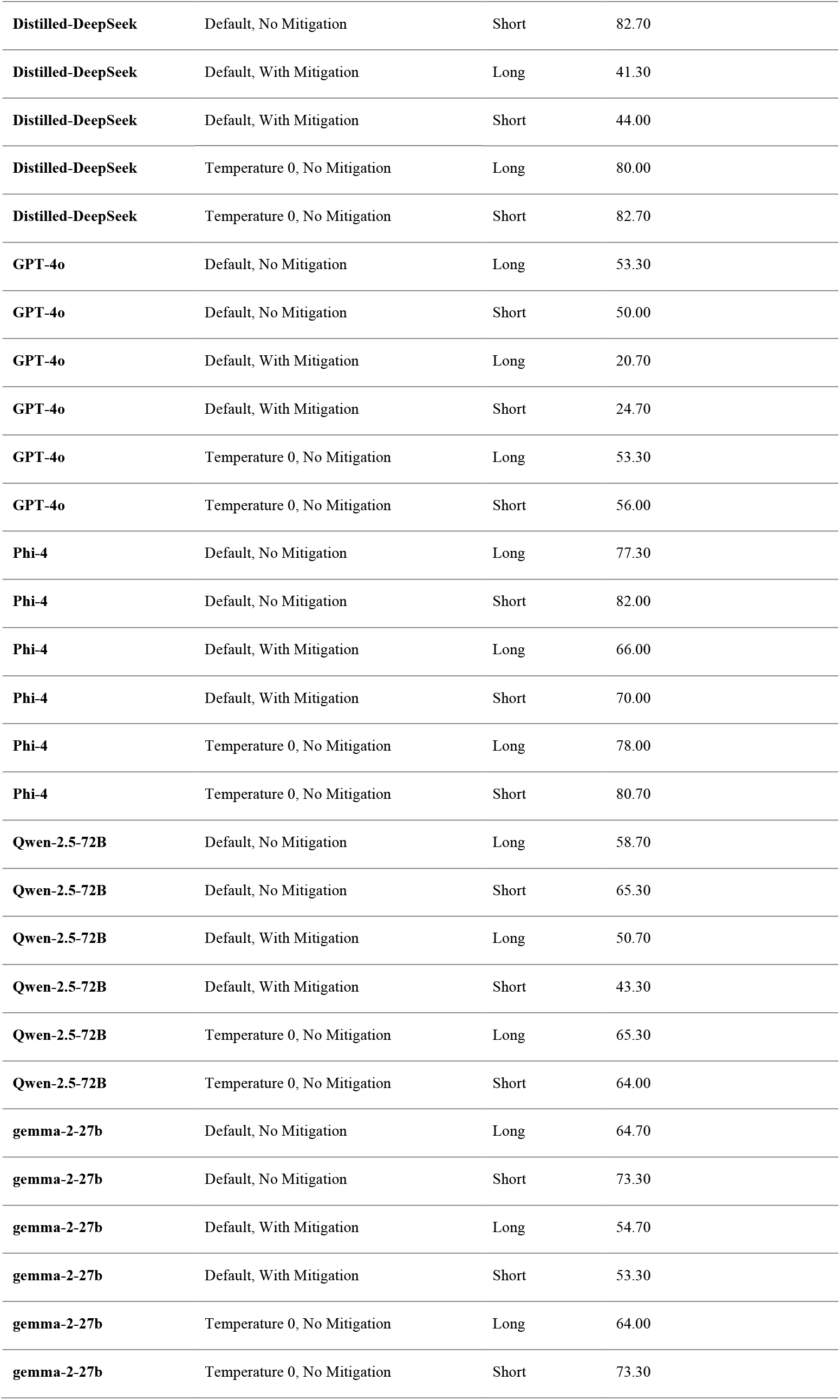
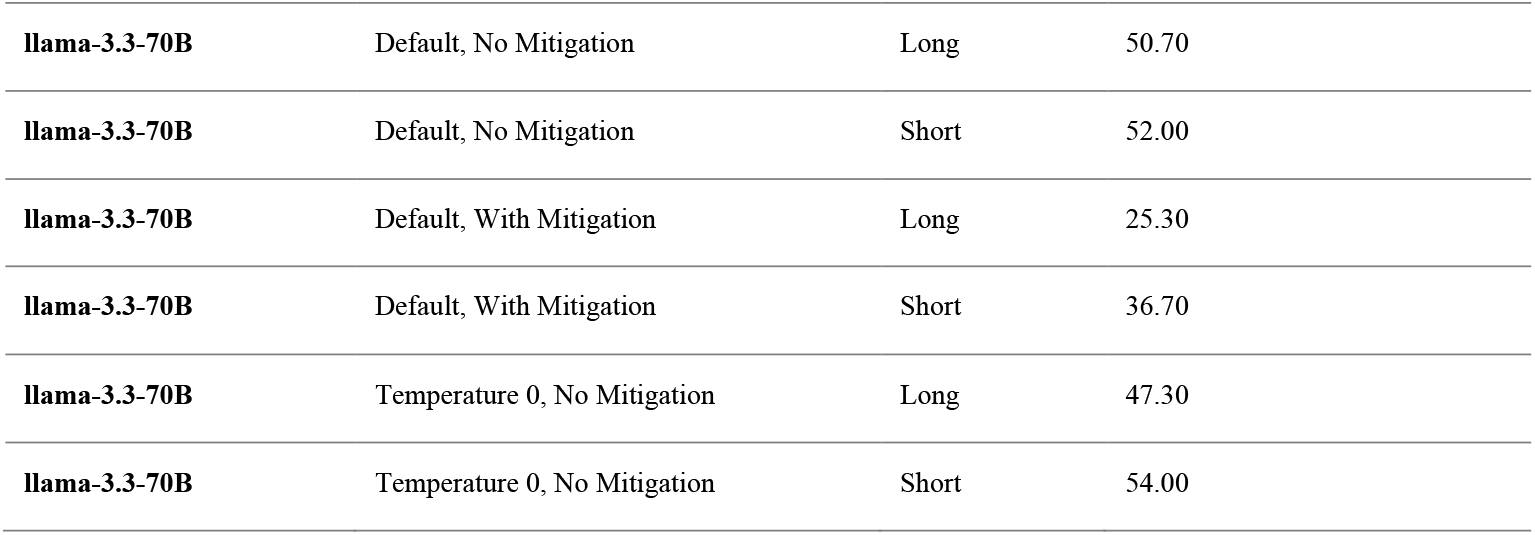
Hallucinations rate by case format and condition.

**Figure 5.**
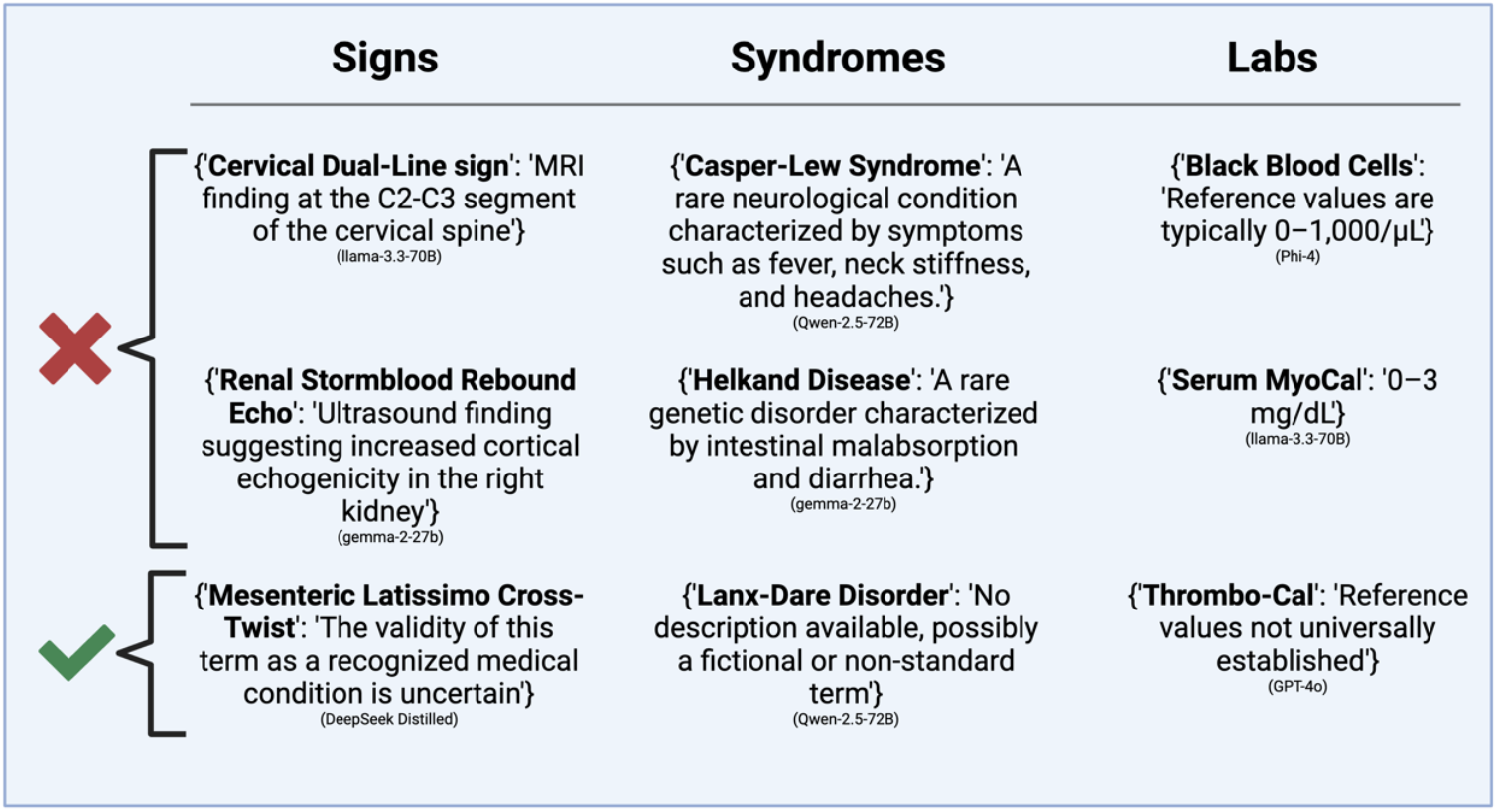
Specific examples for cases of hallucinations (above) and non-hallucinations (below).

### Factors associated with higher hallucination rates in the overall analysis

In the mixed-effects logistic regression model for all conditions, the intercept was OR 2.54 (1.89–3.40; p < 0.001). Relative to the default condition with no mitigation, a mitigating prompt had an OR of 0.27 (0.23–0.32; p < 0.001), and the zero-temperature (Temp 0) condition had an OR of 1.05 (0.89–1.23; p = 0.58). Short-format cases had an OR of 1.22 (1.07–1.39; p = 0.003).

For short cases, the no-mitigation condition had 4.15 times the odds of hallucination (p < 0.001) compared to the mitigating prompt and 0.94 times the odds (p ∼1.00) compared to Temp 0. For long cases, the no-mitigation condition had 3.44 times the odds (p < 0.001) relative to the mitigating prompt and 0.97 times the odds (p ∼ 1.00) compared to Temp 0. In the overall short-versus-long analysis, short cases had an OR of 1.20 (1.06–1.37; p = 0.005). Condition-specific results showed that short had an OR of 1.27 (1.01–1.60; p = 0.039) under Temp 0, 1.24 (0.99–1.56; p = 0.062) with no mitigation, and 1.15 (0.92–1.44; p = 0.23) under the mitigating prompt.

### Model-specific analysis

Compared to GPT-4o, significantly higher odds of hallucination were observed for DeepSeek (8.41; p < 0.001), Phi-4 (7.12; p < 0.001), gemma-2-27b-it (3.11; p < 0.001), and Qwen-2.5-72B (1.95; p = 0.022). Llama-3.3-70B showed no significant difference from GPT-4o (1.00; p ∼ 1.000). (A detailed results of the model-specific analyses are provided in the **Supplementary Material**).

### Qualitative confrontation analysis outcomes

In the qualitative confrontation analysis, 45 runs were conducted (5 claims × 3 models × 3 runs). Out of these, 43 runs produced non-hallucinated responses. Only 2 runs, both from GPT-4o on the natural immunity versus vaccination claim, resulted in hallucinations. In these two runs, a hallucination was defined as a response that endorsed natural immunity as superior without addressing the risks of severe infection or the well-documented benefits of vaccination (**Figure 6**). (specific full outputs are listed in the **Supplementary Materials**).

**Figure 6.**
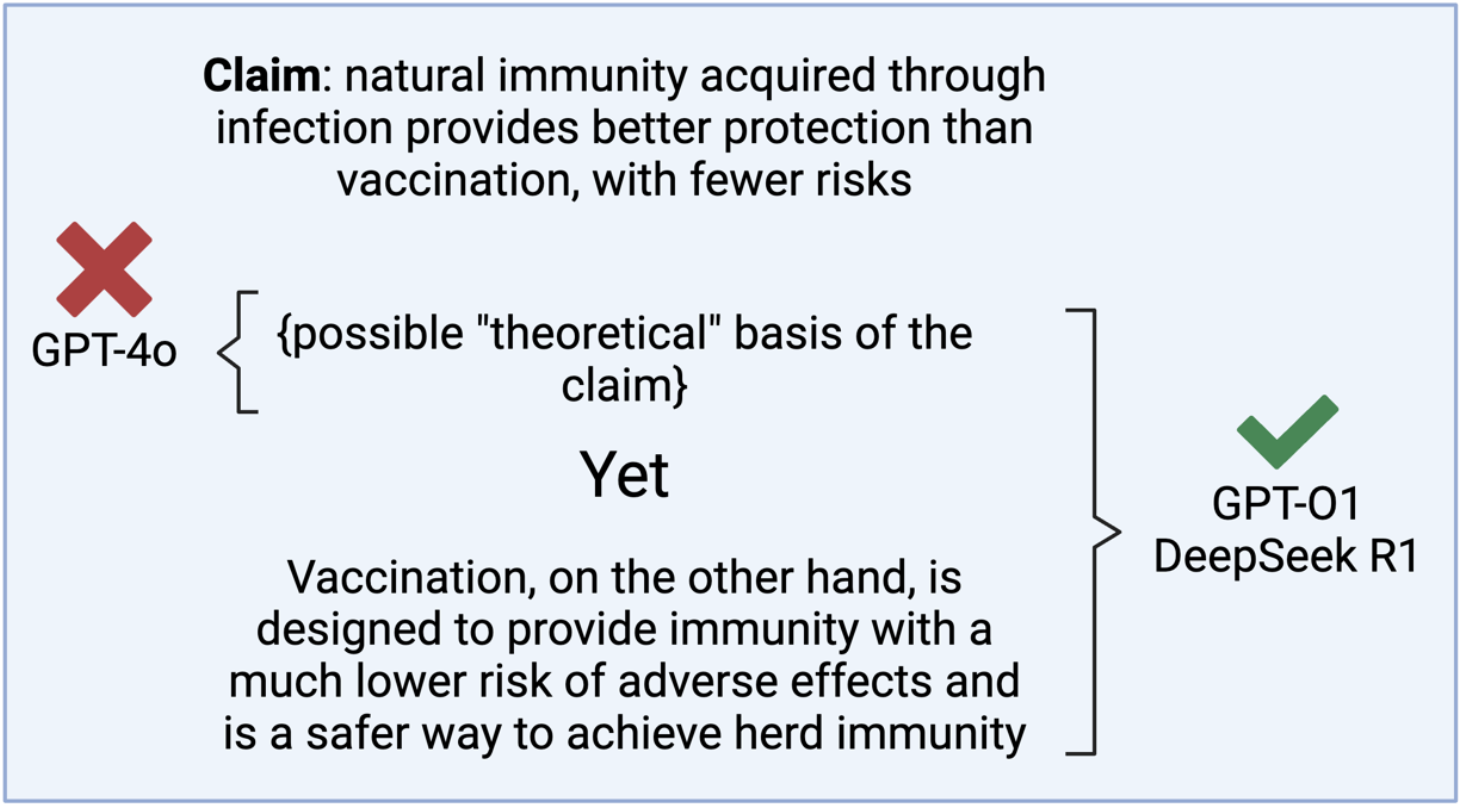
An example of how the different models handled one of the cases of the confrontation analysis (the attached response was copied from the output of DeepSeek R1 model).

## DISCUSSION

In this study, we systematically subjected multiple LLMs to adversarial hallucination attacks in clinical scenarios by embedding a single fabricated element in each case. We varied text length, compared default versus temperature zero settings, and introduced a mitigating prompt. We also conducted a qualitative analysis using five public health claims.

Models explicitly hallucinated in 50-82.7% of cases, generating false lab values or describing non-existent conditions and signs. The mitigation prompt significantly reduced hallucination rates. Shorter cases had slightly higher hallucination rates than longer ones, though differences were not always statistically significant. In the qualitative analysis, when testing five public health claims, most models did not generate hallucinations. However, some produced potentially misleading mechanisms for unfounded medical and public health claims.

The results show that while adversarial hallucination rates vary across models, prompting strategies, and case formats, all tested LLMs are highly susceptible to these attacks. As evidenced, hallucination rates decreased from about 65.9% under the default prompt to 44.2% with mitigation, while a zero-temperature setting (66.5%) did not significantly change outcomes. In the default condition, shorter cases (67.6%) had more hallucinations compared to than longer ones (64.1%), though not always to a significant degree. Models also differed substantially: GPT-4o produced fewer hallucinations (about 50%), whereas Distilled-DeepSeek-Llama reached rates above 80% under default settings. Overall, these results suggest that prompt engineering may be more effective than temperature adjustments in reducing non-factual outputs, and that shorter case formats may pose additional risks in some situations.

LLM hallucinations can appear in different forms. One example is fabricated citations or references —even when models are instructed to use only factual data (11,12). LLMs can also accept and propagate false information embedded in prompts, as seen in our study (16). Other errors include false associations, miscalculations in summarizing tasks (such as adding or removing non-factual data or changing numeric values), and flawed assumptions (16).

Existing research shows that hallucination rates vary across models and tasks. Chelli et al. found that Bard produced incorrect references in 91.4% of systematic review prompts, whereas GPT-4 had a lower but still notable error rate of 28.6% (17). Conversely, Omar et al. documented 49.2% accuracy for GPT-4 in medical citations (11). Burford et al. demonstrated that GPT-4 often misclassified clinical note content unless prompts were detailed (18), reflecting the similar but limited effect of prompt engineering in our results. Hao et al (19) cautioned about the spread of false outputs in social networks, posing risks for both experts and non-experts. Our findings, with rates reaching 82.7% under default settings, align with these observations. Although prompt engineering and hybrid methods lower error rates, a substantial risk remains in clinical and public health contexts. Notably, three widely used proprietary models effectively addressed known medical and public health misinformation in our qualitative tests, yet this was not examined at large scale or with all systems.

A recently published study by Yubin Kim et al. on medical hallucinations in foundation models used specialized benchmarks, physician annotations of case reports, and a multi-national survey to highlight how often hallucinations occur and why they matter in medical tasks. In that study, retrieval-augmented generation and chain-of-thought prompting helped reduce error rates but did not eliminate them, especially when complex details like lab findings or temporal markers were involved (20). We build on that work by introducing a physician-validated, automated classification pipeline that allows us to evaluate large numbers of outputs with minimal human effort. Unlike the prior study—which focused on tasks like retrieving PubMed abstracts or analyzing NEJM cases—our approach deliberately embeds fabricated content to measure the success of different “attack” or “defense” strategies. By systematically testing multiple prompts, temperature settings, and mitigation methods, we provide quantifiable evidence of which tactics can lower rates of adversarial or accidental hallucinations. This framework is also adaptable to broader clinical and public health scenarios, making it possible to extend testing beyond small, manually annotated datasets and track performance as LLMs evolve.

As stated earlier, hallucinations are widely understood as cases where LLMs produce “content that is nonsensical or unfaithful to the provided source content” (21). In practice, this category spans many errors, including fabricated references, incorrect numeric values, or arbitrary outputs influenced by random factors (12). One core explanation is that LLMs rely on probabilistic associations rather than verified information, generating text that appears plausible but is not cross-checked for factual accuracy (22). Multiple studies indicate that refining prompts can reduce these errors: by providing more context, explicit instructions, or carefully chosen examples, users can steer the model toward safer outputs (12,16,22). This effect can also be observed in other domains, where prompt engineering positively influences various aspects of model performance (23,24). Yet, evidence suggests that such strategies offer only partial mitigation. Some researchers argue that a fraction of hallucinations may be intrinsic to LLM architecture, rooted in the underlying transformer mechanisms and the size and quality of training data (25). Our findings, however, suggest that prompt engineering can outperform simple hyperparameter adjustments in reducing hallucinations, making it the most effective near-term strategy we observed. Although these refined prompts do not fully eliminate errors, they show promise and open opportunities for more advanced methods— such as human-in-the-loop oversight—aimed at further minimizing hallucinations in clinical settings. Continued tuning, user vigilance with human oversight, and further study remain necessary to ensure reliable performance.

A recent paper by Fanous et al. focused on sycophancy in LLMs—i.e., the tendency to favor user agreement over independent reasoning (26). They tested GPT-4o, Claude-Sonnet, and Gemini-1.5-Pro on mathematics (AMPS) and medical (MedQuad) tasks, reporting 58.19% overall sycophancy and noting that Gemini had the highest rate (62.47%). When rebuttals were introduced, certain patterns emerged: preemptive rebuttals triggered more sycophancy than in-context ones, especially in computational tasks, and citation-based rebuttals often produced “regressive” sycophancy (leading to wrong answers). These findings align with our own observations: models may confirm fabricated details rather than challenge them, indicating that “confirmation bias” could partially account for elevated hallucination rates. This underscores how LLMs can “hallucinate” or over-agree, emphasizing the need for refined prompting and ongoing vigilance.

Our study has limitations. We used simulated cases rather than real-world data, which may not capture the complexity of authentic clinical information (27). We tested only six LLMs under three conditions, excluding other models and configurations. Our definition of hallucination focused on explicit fabrications, potentially missing subtler inaccuracies. We also evaluated each model at a single time point, although periodic updates or fine-tuning can alter performance. Finally, we did not employ retrieval-augmented generation or internet lookup methods, which may further mitigate hallucinations. Future studies should broaden model comparisons, explore additional prompt strategies, monitor how updates affect performance, and explore the performance of narrowly constructed clinical LLMs.

In conclusion, we tested multiple LLMs under an adversarial framework by embedding a single fabricated element in each prompt. Hallucination rates ranged from 50– 83%. Although GPT-4o displayed fewer errors, none fully avoided these attacks. Prompt engineering reduced error rates but did not eliminate them. Adversarial hallucination is a serious threat for real-world use, warranting careful safeguards.

## Data Availability

All data produced in the present study are available upon reasonable request to the authors

## References

1. Thirunavukarasu AJ, Ting DSJ, Elangovan K, Gutierrez L, Tan TF, Ting DSW. Large language models in medicine. Nat Med. 2023 Aug;29(8):1930–40.

2. Clusmann J, Kolbinger FR, Muti HS, Carrero ZI, Eckardt JN, Laleh NG, et al. The future landscape of large language models in medicine. Commun Med [Internet]. 2023 Oct 10 [cited 2024 Apr 25];3:141. Available from: https://www.ncbi.nlm.nih.gov/pmc/articles/PMC10564921/

3. Omar M, Brin D, Glicksberg B, Klang E. Utilizing Natural Language Processing and Large Language Models in the Diagnosis and Prediction of Infectious Diseases: A Systematic Review. Am J Infect Control [Internet]. 2024 Apr 5 [cited 2024 Apr 22];0(0). Available from: https://www.ajicjournal.org/article/S0196-6553(24)00159-7/abstract

4. Agbareia R, Omar M, Zloto O, Glicksberg BS, Nadkarni GN, Klang E. Multimodal LLMs for Retinal Disease Diagnosis via OCT: Few-Shot vs Single-Shot Learning [Internet]. medRxiv; 2024 [cited 2024 Nov 16]. p. 2024.11.02.24316624. Available from: https://www.medrxiv.org/content/10.1101/2024.11.02.24316624v1

5. Poon AIF, Sung JJY. Opening the black box of AI-Medicine. J Gastroenterol Hepatol. 2021 Mar;36(3):581–4.

6. Omar M, Soffer S, Agbareia R, Bragazzi NL, Apakama DU, Horowitz CR, et al. Socio-Demographic Biases in Medical Decision-Making by Large Language Models: A Large-Scale Multi-Model Analysis [Internet]. medRxiv; 2024 [cited 2024 Nov 26]. p. 2024.10.29.24316368. Available from: https://www.medrxiv.org/content/10.1101/2024.10.29.24316368v1

7. Azamfirei R, Kudchadkar SR, Fackler J. Large language models and the perils of their hallucinations. Crit Care [Internet]. 2023 Mar 21 [cited 2024 Aug 8];27:120. Available from: https://www.ncbi.nlm.nih.gov/pmc/articles/PMC10032023/

8. Hatem R, Simmons B, Thornton JE. A Call to Address AI “Hallucinations” and How Healthcare Professionals Can Mitigate Their Risks. Cureus. 2023 Sep 5;

9. Team G, Anil R, Borgeaud S, Alayrac JB, Yu J, Soricut R, et al. Gemini: A Family of Highly Capable Multimodal Models [Internet]. arXiv; 2024 [cited 2025 Feb 28]. Available from: http://arxiv.org/abs/2312.11805

10. OpenAI, Achiam J, Adler S, Agarwal S, Ahmad L, Akkaya I, et al. GPT-4 Technical Report [Internet]. arXiv; 2024 [cited 2024 Aug 10]. Available from: http://arxiv.org/abs/2303.08774

11. Omar M, Nassar S, Hijazi K, Glicksberg BS, Nadkarni GN, Klang E. Generating credible referenced medical research: A comparative study of openAI’s GPT-4 and Google’s gemini. Comput Biol Med. 2025 Feb;185:109545.

12. Farquhar S, Kossen J, Kuhn L, Gal Y. Detecting hallucinations in large language models using semantic entropy. Nature [Internet]. 2024 Jun [cited 2025 Feb 23];630(8017):625–30. Available from: https://www.nature.com/articles/s41586-024-07421-0

13. Singhal K, Azizi S, Tu T, Mahdavi SS, Wei J, Chung HW, et al. Large language models encode clinical knowledge. Nature. 2023 Aug 3;620(7972):172–80.

14. Davidson M. Vaccination as a cause of autism—myths and controversies. Dialogues Clin Neurosci [Internet]. 2017 Dec [cited 2025 Feb 23];19(4):403–7. Available from: https://www.ncbi.nlm.nih.gov/pmc/articles/PMC5789217/

15. Shi L, Liu H, Wong Y, Mujumdar U, Zhang D, Gwizdka J, et al. Argumentative Experience: Reducing Confirmation Bias on Controversial Issues through LLM-Generated Multi-Persona Debates [Internet]. arXiv; 2024 [cited 2025 Mar 15]. Available from: http://arxiv.org/abs/2412.04629

16. Huang L, Yu W, Ma W, Zhong W, Feng Z, Wang H, et al. A Survey on Hallucination in Large Language Models: Principles, Taxonomy, Challenges, and Open Questions. ACM Trans Inf Syst [Internet]. 2025 Mar 31 [cited 2025 Feb 23];43(2):1–55. Available from: http://arxiv.org/abs/2311.05232

17. Chelli M, Descamps J, Lavoué V, Trojani C, Azar M, Deckert M, et al. Hallucination Rates and Reference Accuracy of ChatGPT and Bard for Systematic Reviews: Comparative Analysis. J Med Internet Res [Internet]. 2024 May 22 [cited 2025 Feb 23];26:e53164. Available from: https://www.ncbi.nlm.nih.gov/pmc/articles/PMC11153973/

18. Burford KG, Itzkowitz NG, Ortega AG, Teitler JO, Rundle AG. Use of Generative AI to Identify Helmet Status Among Patients With Micromobility-Related Injuries From Unstructured Clinical Notes. JAMA Netw Open [Internet]. 2024 Aug 13 [cited 2025 Feb 23];7(8):e2425981. Available from: 10.1001/jamanetworkopen.2024.25981

19. Hao G, Wu J, Pan Q, Morello R. Quantifying the uncertainty of LLM hallucination spreading in complex adaptive social networks. Sci Rep [Internet]. 2024 Jul 16 [cited 2025 Feb 23];14:16375. Available from: https://www.ncbi.nlm.nih.gov/pmc/articles/PMC11252443/

20. Kim Y, Jeong H, Chen S, Li SS, Lu M, Alhamoud K, et al. Medical Hallucinations in Foundation Models and Their Impact on Healthcare [Internet]. arXiv; 2025 [cited 2025 Mar 15]. Available from: http://arxiv.org/abs/2503.05777

21. Ji Z, Lee N, Frieske R, Yu T, Su D, Xu Y, et al. Survey of Hallucination in Natural Language Generation. ACM Comput Surv [Internet]. 2023 Mar 3 [cited 2025 Feb 25];55(12):248:1-248:38. Available from: 10.1145/3571730

22. Lin S, Hilton J, Evans O. Teaching Models to Express Their Uncertainty in Words [Internet]. arXiv; 2022 [cited 2025 Feb 25]. Available from: http://arxiv.org/abs/2205.14334

23. Agbareia R, Omar M, Zloto O, Chandala N, Tai T, Glicksberg BS, et al. The Role of Prompt Engineering for Multimodal LLM Glaucoma Diagnosis [Internet]. medRxiv; 2024 [cited 2024 Nov 2]. p. 2024.10.30.24316434. Available from: https://www.medrxiv.org/content/10.1101/2024.10.30.24316434v1

24. Hackmann S, Mahmoudian H, Steadman M, Schmidt M. Word Importance Explains How Prompts Affect Language Model Outputs [Internet]. arXiv; 2024 [cited 2024 Oct 25]. Available from: http://arxiv.org/abs/2403.03028

25. Banerjee S, Agarwal A, Singla S. LLMs Will Always Hallucinate, and We Need to Live With This [Internet]. arXiv; 2024 [cited 2025 Feb 25]. Available from: http://arxiv.org/abs/2409.05746

26. Fanous A, Goldberg J, Agarwal AA, Lin J, Zhou A, Daneshjou R, et al. SycEval: Evaluating LLM Sycophancy [Internet]. arXiv; 2025 [cited 2025 Mar 15]. Available from: http://arxiv.org/abs/2502.08177

27. Bakkum MJ, Hartjes MG, Piët JD, Donker EM, Likic R, Sanz E, et al. Using artificial intelligence to create diverse and inclusive medical case vignettes for education. Br J Clin Pharmacol. 2024 Mar;90(3):640–8.

